# A serum protein network predicts the need for systemic immunomodulatory therapy in autoimmune uveitis

**DOI:** 10.1101/2021.09.22.21263286

**Authors:** Jonas J.W. Kuiper, Fleurieke H. Verhagen, Sanne. Hiddingh, Roos A.W. Wennink, Anna M. Hansen, Kerry A. Casey, Imo E. Hoefer, Saskia Haitjema, Julia Drylewicz, Mehmet Yakin, H. Nida Sen, Timothy R.D.J. Radstake, Joke H. de Boer

## Abstract

Objective biomarkers that can predict a severe disease course of autoimmune uveitis are lacking, and warranted for early identification of high-risk patients to improve visual outcome. The need for non-steroid immunomodulatory therapy (IMT) to control autoimmune uveitis is indicative of a more severe disease course. We used aptamer-based proteomics and a bioinformatic pipeline to uncover the serum protein network of 52 treatment-free patients and 26 healthy controls, and validation cohorts of 114 and 67 patients. Network-based analyses identified a highly co-expressed serum signature (n=85 proteins) whose concentration was consistently low in controls, but varied between cases. Patients that were positive for the signature at baseline showed a significantly increased risk for IMT during follow-up, independent of anatomical location of disease. In an independent cohort (n=114), we established robust risk categories that confirmed that patients with high levels of the signature at diagnosis had a significantly increased risk to start IMT during follow-up. Finally, we further validated the predictive power of the signature in a third cohort of 67 treatment-naive North-American patients. A serum protein signature was highly predictive for IMT in human autoimmune uveitis and may serve as an objective blood biomarker to aid in clinical-decision making.

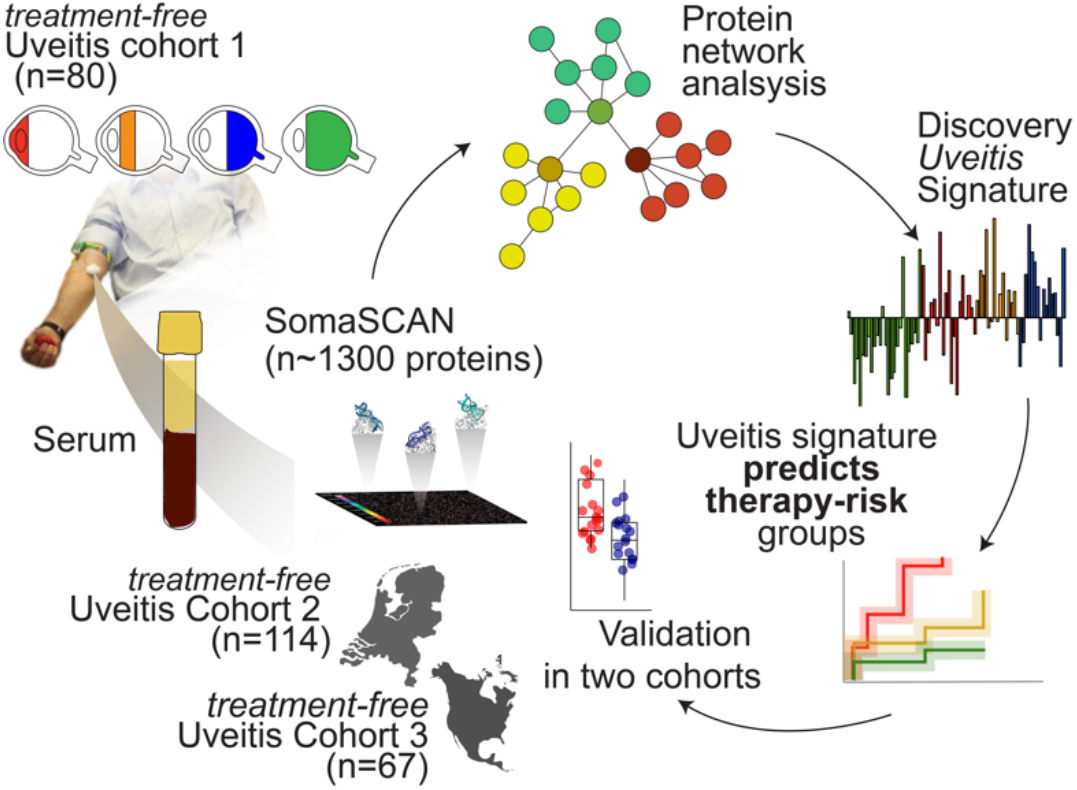

## Introduction

Autoimmune or non-infectious uveitis is a spectrum of severe inflammatory disorders of the inner eye with complex inflammatory etiologic origins that often cause decreased vision or in some cases blindness. Vision loss as a result of inflammation and its complications can be limited or reversed by early and adequate therapy (1). Local or systemic corticosteroid treatment are the first-line therapy for uveitis, but steroid-sparing agents are recommended when inflammation cannot be controlled by corticosteroids or to limit steroid side-effects in cases where long-term treatment is required (2).

Non-steroid immunomodulatory therapy (IMT, including antimetabolites, calcineurin inhibitors, alkylating agents, and biologicals) is very effective in preventing vision loss but because of potential adverse effects, it is typically reserved for severe vision threatening uveitis (3, 4). In general, available strategies that aid in the identification of patients that require IMT are based on establishing the severity of uveitis *ad hoc* or by imaging the affected ocular tissues. Minimally invasive molecular tools to objectively predict a severe disease course (far) in advance are lacking but are highly warranted to help better identify those patients who are at risk and will need IMT to control their disease (5). The blood proteomic fingerprint of non-infectious uveitis patients may identify immune signatures that can be exploited to stratify high risk patients during diagnostic or periodic work-up to allow prompt clinical decision making to prevent poor visual outcome.

## Results and Discussion

We used *SomaScan* aptamer technology to measure 1,305 serum proteins in 54 treatment-free uveitis patients with active uveitis (**Table 1**) and 26 healthy individuals (**Figure 1A**). A link to the full reproducible code and raw data used in this study can be found in the *Methods*. After quality control, two outlier samples were removed (**Figure 1B**). We detected 936 serum proteins of which 193 were differentially expressed (DEPs) between the disease groups (*likelihood ratio test*, false discovery rate of 5%) (**Supplemental Table 1**). Global comparisons by hierarchical cluster analysis discerned three clusters of differentially expressed proteins (Cl1, Cl2 and Cl3) and two large clusters of samples; cluster A containing most controls (23/26 controls are in this cluster) and 11 patients, while cluster B contained nearly exclusively patients (41/44) (**Figure 1C**). Protein cluster Cl1 contained proteins that were higher in concentration in serum of uveitis patients, including *S100A12*, and *Annexin A1* (**Figure 1C**), and was most enriched for the *Neutrophil Degranulation* pathway (*Padj* = 1.6×10^−21^)(**Figure 1C**). The levels of proteins of cluster Cl2 were generally lower in serum of patients compared to controls (e.g., Interferon Beta 1), while proteins of cluster Cl3 often showed uveitis subtype-specific expression patterns (e.g., ERAP1 in anterior uveitis) (**Figure 1C**).

**Table 1.** Demographic details of patients in the three cohorts investigated in this study. Details are provided per anatomical location.

| Patient cohort | Cohort 1(n=52),<br>Netherlands | Cohort 2 (n=114),<br>Netherlands | Cohort 3 (n=67),<br>U.S.A. |
| --- | --- | --- | --- |
| Anatomical location |  |  |  |
| *Anterior (%)* | 19 (37) | 36(31.5) | - |
| Female/Male | 14/5 | 22/14 | - |
| Mean Age (SD) | 47(16) | 45(19) | - |
| *Intermediate (%)* | 15 (29) | 9 (8) | 25 (37.3) |
| Female/Male | 10/5 | 5/4 | 14/11 |
| Mean Age (SD) | 37(12) | 40(21) | 35(18) |
| *Posterior (%)* | 18 (34) | 25(22) | 15(22.4) |
| Female/Male | 9/9 | 15/10 | 15/0 |
| Mean Age (SD) | 52(12) | 50(18) | 51(15) |
| *Pan (%)* | - | 44(38.5) | 27(40.3) |
| Female/Male | - | 21/23 | 15/12 |
| Mean Age (SD) | - | 44(19) | 41(16) |

**Figure 1.**
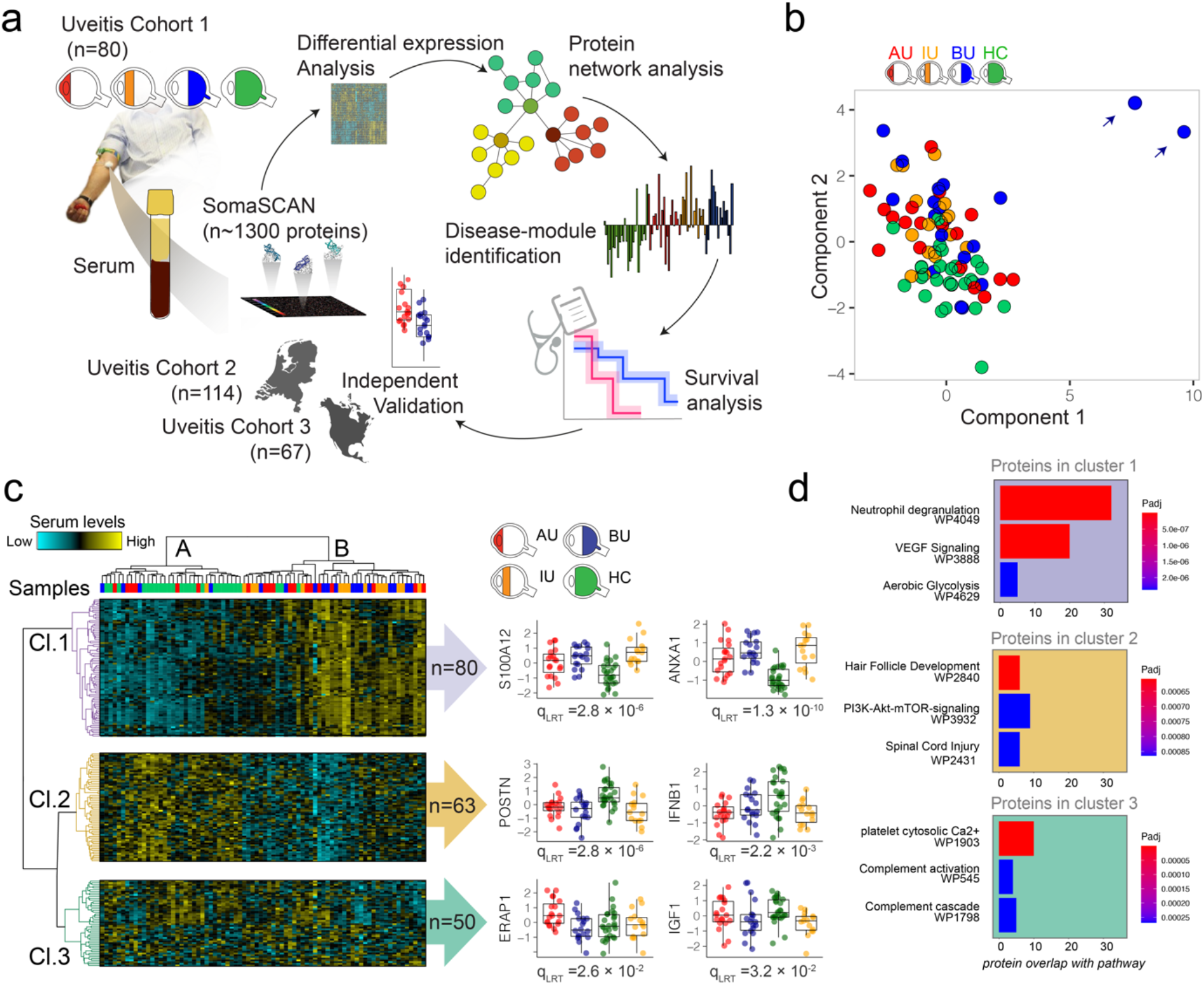
Serum proteome changes in patients with autoimmune uveitis. **a**) Schematic overview of the design of the study. **b**) Principal component analysis based on the log10 transformed relative fluorescence units [RFU] of 938 detected serum proteins in 54 patients with anterior [AU], intermediate [IU], or Birdshot uveitis [BU], and 26 controls. The blue arrows indicate two BU outlier patients removed from further analysis. **c**) Hierarchical cluster analysis (using Euclidean distance with Ward’s minimum variance method) of 193 differentially expressed serum proteins (likelihood ratio test [LRT] q-value<0.05). Three overarching clusters of differentially expressed proteins (rows) are color-coded. Scatterplots of representative serum proteins for each cluster are shown with their respective *q*-values from the LRT. **d**) Top 3 WikiPathways for the differentially expressed proteins in each cluster. IGF1; Insulin-like growth factor 1, ERAP1; Endoplasmic Reticulum Aminopeptidase 1, S100A12; S100 calcium-binding protein A12, ANXA1; Annexin A1, POSTN; Periostin, IFNB1; Interferon Beta 1.

The human serum proteome functions as a biological network with structured co-regulated clusters of proteins (i.e., pathways, cells) (6). We constructed a signed weighted co-expression network that divided the serum proteome (n=936) into nine highly structured protein modules (**Figure 2A** and **Supplemental Table 1)**, ranging in size from 14 to 223 proteins, whereas 37% of detected serum proteins fell outside of these modules (assigned to a ‘grey’ module) (**Figure 2B**). We noted that differentially expressed proteins were proportionally highly prevalent in the blue module (61/193 DEPs of in total 85 proteins in the blue module) (**Figure 2B**). Proteins in the blue module were strongly enriched for the *Neutrophil Degranulation pathway* (*Padj* = 1.2×10^−11^) and included many DEPs from cluster 1 (**Figure 2C**). Among these were S100A12, IMPDH1, and ARG1 that showed high module membership (MM) scores [MM>0.9], further supporting that this module’s is characterized by neutrophil related proteins (**Figure 2C**). Note that the blue module identified here was remarkably similar to a previously identified (6) serum protein module [termed “PM16”] (**Figure 2D**).

**Figure 2.**
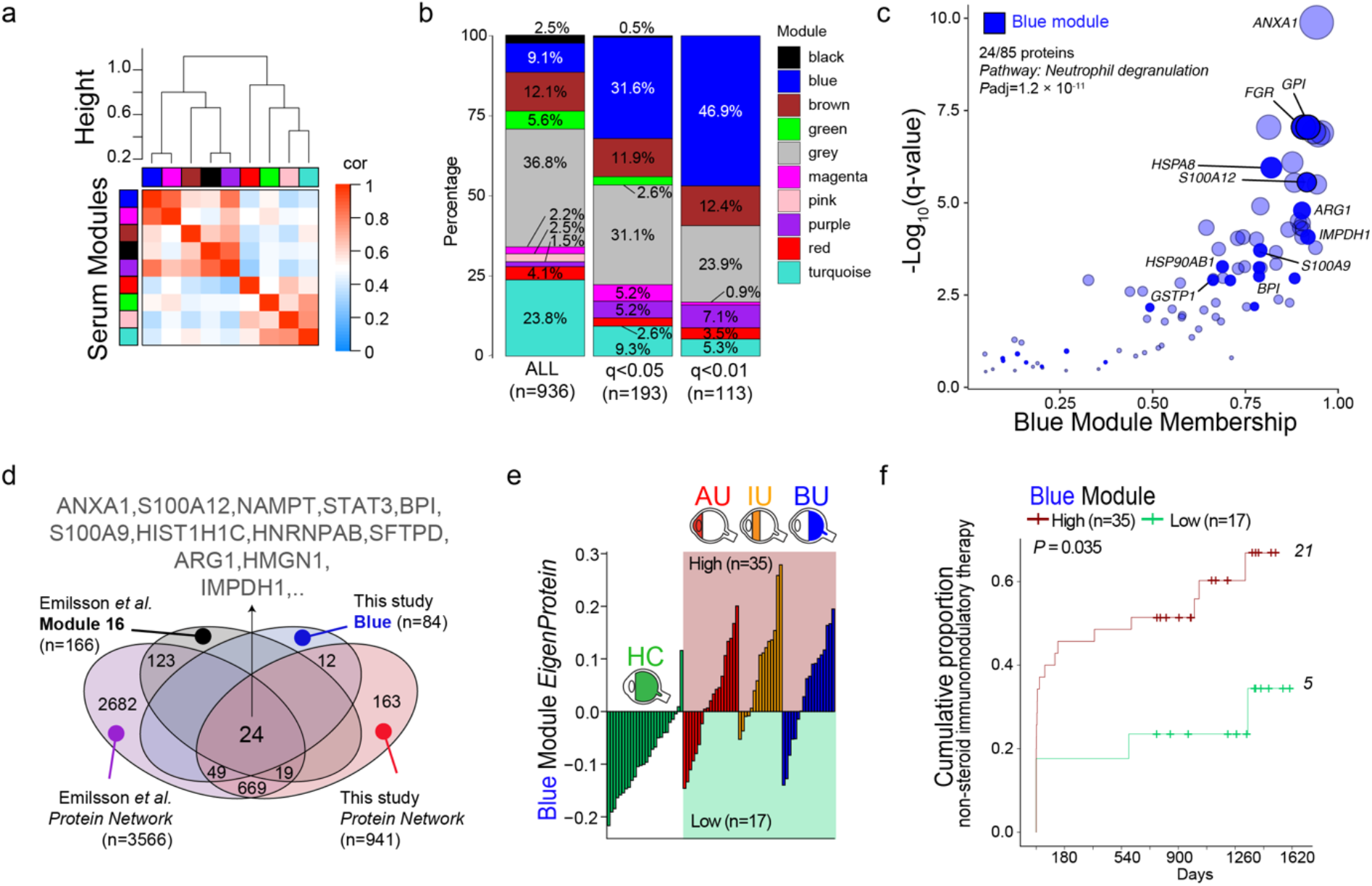
Co-expression network analysis links serum protein-network to systemic immunomodulatory therapy. **a**) Weighted protein co-expression network analysis of 936 proteins discerned 9 (color-coded) serum protein modules. The correlation of the module’s *EigenProtein* is color-coded from blue to red. The correlation (1-cor[*EigenProteins*]) was used as a distance metric for the dendrogram. **b**) The proportion of all detected serum proteins and differentially expressed proteins (at q<0.05 and q<0.01) among the 9 modules identified under *a*. Note that the grey “module” contains unassigned proteins. **c**) The *q*-values from the likelihood ratio test versus the module membership for proteins of the blue module. The size of the circles is proportional to -Log_10_(q-value). 24 (indicated in solid blue) are present in the *neutrophil degranulation* pathway (adjusted *P* value from enrichment analysis) **d**) Venn diagram comparing the serum protein network and protein module 16 of *Emilsson et al. 2018* (6) and the blue module dentified in this study. **e**) The EigenProtein value of the blue module (first principal component of the module) for each control (green) and anterior [AU, red], intermediate [IU, orange], or Birdshot uveitis [BU, blue] patients; 35 patients showed a relatively high expression (‘high’ group) and 17 patients displayed a relatively low expression of the proteins (‘low’ group). **f**) The cumulative event curve for the use of systemic non-steroid immunomodulatory therapy in patients with high- (in red) or low (in green) expression of the blue protein as identified in *e*. The *P* value from a log-rank test and the total IMT events during follow-up per group are shown.

The *eigenProtein* of the blue module (i.e., first principal component of the expression data of this module) was relatively low in controls, but varied among patients regardless of anatomical location of uveitis (**Figure 2E**); 35 patients (67%) showed relatively high levels and 17 patients (33%) displayed relatively low levels of the blue module. Since the two groups of patients stratified by the blue module were highly comparable in age, sex, and anatomical location of disease (**Supplemental Table 2**), we hypothesized that the serum protein module represented immune activation that drives disease severity. Since (non-corticosteroid) *systemic immunomodulatory therapy* (IMT) is often introduced to control severe inflammation (2), we assessed if the blue module could stratify patients into differential risk groups for their need for IMT using survival analysis. The log rank test supported that patients stratified upon the blue module differ significantly in their probability to start IMT during follow-up (*P* = 0.035). Using a multivariate Cox analysis considering age, sex, and anatomical location of uveitis, we identified, as expected, a strong relationship between anatomical location and use of IMT during follow-up (AU versus BU, hazard ratio [HR]=5.5, 95% confidence interval [95%CI]=1.8-16.8, *P* = 0.003)(**Supplemental Figure 2**). Adjusted for anatomical location, age and sex, we confirm that cases with a relatively high expression of the blue module at baseline have a higher risk for requiring IMT (HR [95%CI] = 3.42[1.22-9.5], *P* = 0.019). We conclude that the relative levels of a network of serum proteins at baseline can distinguish patients with a differential risk for systemic immunosuppressive therapy.

Next, we sought to independently validate the association between the blue module and the risk for IMT. Because the module was strongly enriched for neutrophil function, we assessed if key proteins from this module were indeed expressed in neutrophils. To this end, we compared the levels of these proteins in proteomic data from 27 primary blood immune cell subsets. This analysis showed that many proteins from the blue module, such as S100A12, and Annexin A1 were specifically highly expressed in Neutrophils (**Figure 3A**, and **Supplemental Table 3**). Also, the blue module’s *EigenProtein* correlated significantly with the neutrophil count in blood (r = 0.57, *P* = 0.006) (**Figure 3B**) suggesting that the blood neutrophil count could serve as a proxy for the blue module. This would also overcome the limitation of the SOMAscan technology (provides a relative abundance of protein) and help to define objective thresholds for the signature for independent validation.

**Figure 3.**
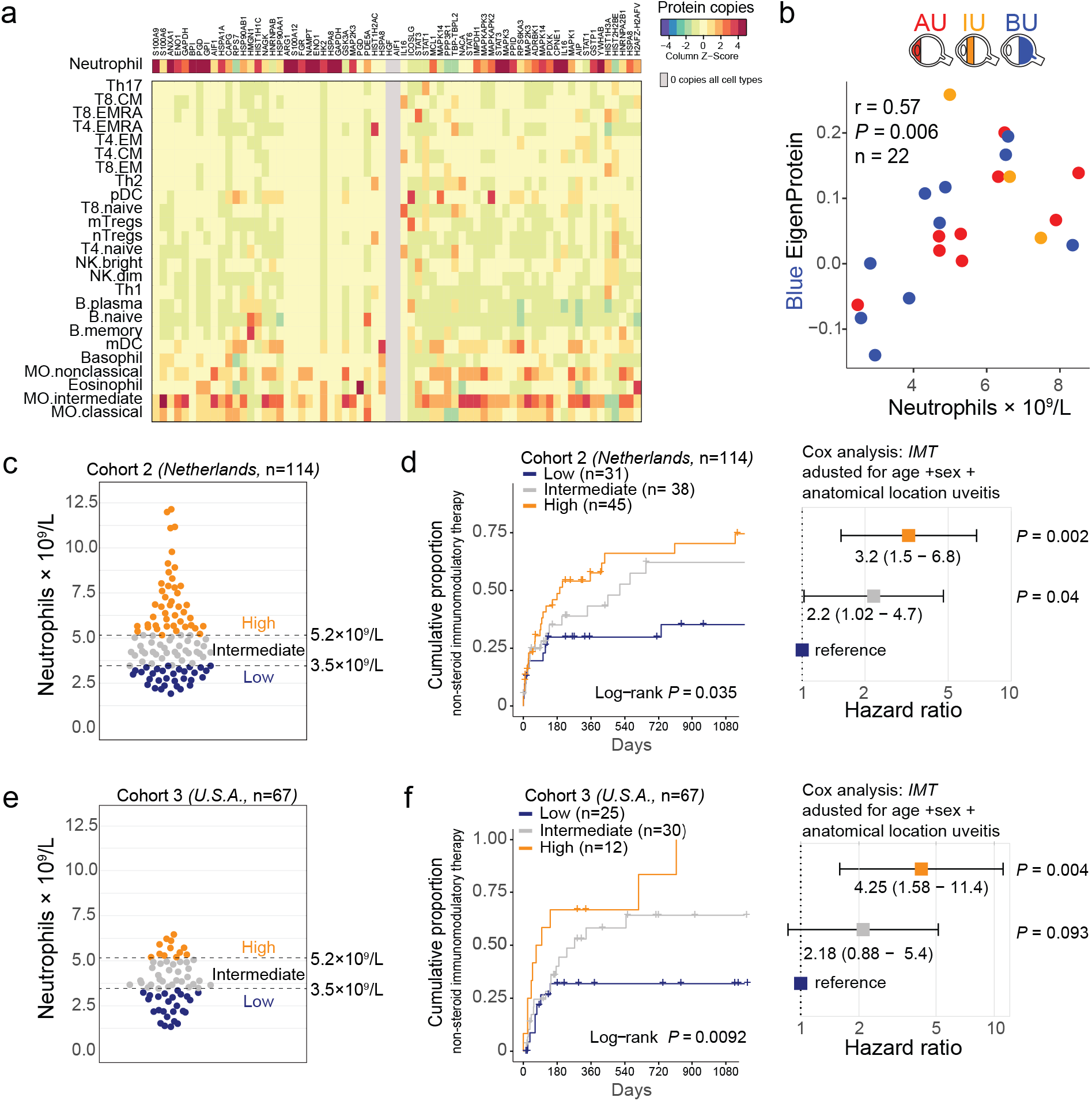
Blood neutrophil count is a proxy for the serum signature and predicts risk of systemic immunomodulatory therapy. **a**) Heatmap of the mean protein copy numbers (Z-score) in primary neutrophils and other immune cell subsets (data from Rieckmann et al.,(7)) for proteins identified in the blue serum protein module. Details on the protein copies per cell type are outlined in Table *E2*. **b**) Scatter plot of the EigenProtein values for the blue module versus the blood neutrophil count for 22 anterior [AU], intermediate [IU], or Birdshot uveitis [BU] cases with available blood neutrophil count data at baseline. The correlation coefficient *r* and *P* value are from Pearson’s product-moment correlation test. **c**) The blood neutrophil count of an independent cohort of Dutch non-infectious uveitis patients. The split points used to stratify the patients into three groups (low, intermediate, and high, respectively) for survival analysis are indicated. **d**) On the left; the cumulative event curve for the use of systemic immunomodulatory therapy in the Dutch cohort stratified by baseline blood neutrophil group. On the right: corresponding forest plot (cox proportional hazard analysis adjusted for age, sex, and anatomical location of uveitis) for the use of systemic immunomodulatory therapy among the ‘low’ (reference), ‘intermediate’, and ‘high’ blood neutrophil groups. **e, f**) Same as in *c* and *d*, but for a cohort of 67 American non-infectious uveitis patients.

We further assessed in an independent cohort (i.e., *cohort 2*, n=114, **Table 1**) of Dutch uveitis patients free of IMT at the time of sampling (i.e., at diagnosis) to validate if the blood neutrophil count was associated with the likelihood for requiring IMT. We calculated optimal split points (see *methods*) in the blood neutrophil count that best stratified cases that required IMT during follow-up from cases that did not, which revealed two major split points at 3.5×10^9^ cells/L and at 5.2×10^9^ cells/L (**Supplemental Figure 2**). We next divided patients into three corresponding categories of baseline blood neutrophil groups; “low” (≤3.5×10^9^/L, n=31), “intermediate” (>3.5×10^9^/L and ≤5.2×10^9^/L, n=38), and “high” (>5.2×10^9^/L, n=45)(**Figure 3C**) and computed hazard functions for these categories. Note that these categories all fall within the normal range for blood neutrophil count. Cox proportional hazard analysis, adjusting for age, sex, and anatomical location of uveitis revealed a >3 times higher risk to start IMT in the “high” group versus the “low” group (HR [95%CI] = 3.2 [1.5-6.8], *P* = 0.002) (**Figure 3D**) (**Supplemental Figure 3**).

To further validate these results, we assessed the relationship between the signature and IMT, using baseline neutrophil count as proxy in a third cohort of 67 treatment naive non-infectious uveitis patients (cohort 3, **Table 1**) from the National Eye Institute in Bethesda, Maryland. Patients from this cohort were divided into the same categories “low” (≤3.5×10^9^/L, n=25), “intermediate” (>3.5×10^9^/L - ≤5.2×10^9^/L, n=30), and “high” (>5.2×10^9^/L, n=12) (**Figure 3E**) and we assessed risk for IMT using cox proportional hazard analysis. This analysis confirmed the significantly higher risk for IMT in the “high” group versus the “low” group (HR= 4.3 [95%CI: 1.58-11.4], *P* = 0.004) **(Figure 3F**) (**Supplemental Figure 4**). Note that iterations of the optimal split points in neutrophil blood count from cohort 3 (measured by another platform, see Online Methods) revealed optimal split points nearly identical to cohort 2 (3.4×10^9^/L and 5.2×10^9^/L) (**Supplemental Figure 2**), supporting that the defined neutrophil categories are clinically robust across patient populations.

Collectively, these results show that the levels of a serum protein network linked to blood neutrophils at baseline (i.e., diagnosis) was highly predictive for the need for IMT during follow-up. Our results revealed that relative thresholds in normal blood neutrophils can serve as a routinely available proxy for the serum signature and could robustly stratify patients into differential risk categories. Alongside ophthalmological work-up and imaging, detection of the signature at disease onset can help to better identify patients with a severe disease course, which can be helpful to both inform the patient about expected disease progression and for developing treatment strategies. Since the blood neutrophil count can be easily monitored during diagnostic work-up, the implementation of the here defined thresholds for prospective evaluation of these results should be possible in most clinical settings, and as we show, across at least two common haematology platforms for quantification of whole blood samples. Since none of the patients were on systemic therapy at the time of sampling, it is currently unknown how our findings are applicable to uveitis patients on IMT, which is a limitation of our study. Future research will focus on careful monitoring of neutrophil count to investigate if it can also predict disease relapse or treatment response, and potentially for making therapeutic decisions to deliver personalised care for patients with intraocular inflammatory diseases.

## Methods

### Patient cohorts

Serum from cohort 1 was collected (July 2014-December 2016) from 54 adult patients with HLA-B27-positive anterior uveitis (AU), idiopathic intermediate uveitis (IU), and HLA-A29-positive Birdshot Uveitis at the department of Ophthalmology of the University Medical Center Utrecht, The Netherlands (**Table 1**). Serum from 26 anonymous blood donors (blood donors University Medical Center Utrecht) with no history of inflammatory eye disease served as controls (HC). At the time of sampling, all patients had active uveitis (new onset or relapse) according to the S*tandardization of Uveitis Nomenclature* (SUN criteria (8). All patients did not use systemic treatment in the last 3 months prior to sampling (except for one patient, ≤10 mg oral prednisolone).

### SOMAscan proteomic assay

Serum tubes were kept for 30 minutes at room temperature, centrifuged at 2000g for 10 minutes at room temperature and stored directly at -80 Celcius. Frozen samples were shipped on dry ice to SomaLogic (Boulder, CO, USA). Serum samples were analysed by SomaLogic using the 1.3K SomaScan assay (1,305 proteins)(9). The samples were run with the mitigation protocol at SomaLogic to control assay interference from potential anti-self-nucleic acid autoantibodies (10). All samples passed quality control (see “SOMAscan quality statement” at DataverseNL, https://doi.org/10.34894/QR1VFZ). SomaScan dataset after hybridization control normalization, median signal normalization, and calibration are presented in *adat* format and used for analysis.

### Construction of the serum protein co-expression network

Weighted gene co-expression network analysis (WGCNA) was conducted using the WGCNA package (13). Using a recommended soft thresholding power of 12 for signed networks and a minimal module size of 10 protein, we generated a network with nearly scale-free topology (*r*2 > 0.9). Throughout the manuscript we replaced the WGCNA term “EigenGene” for “EigenProtein”. Modules with highly similar expression profiles (correlation of *EigenProtein* values ≳0.75) were merged. Data for protein module 16 [PM16] as reported by Emilsson *et al*., (6) were extracted from the supplemental data of the manuscript by Emilsson and co-workers. Protein copy numbers of primary human immune cells were obtained from supplemental data of the manuscript by Rieckmann *et al*. (7).

### Survival analysis of Neutrophil blood count

The neutrophil count of 111 treatment-free Dutch patients (cohort 2) was determined in whole blood samples by the CELL-DYN Sapphire automated haematology analyser (Abbott Diagnostics, Santa Clara, CA, USA) and obtained from the *Utrecht Patient Oriented Database* (UPOD) infrastructure of the University Medical Center Utrecht (14). These patients had active uveitis and blood withdrawal was conducted in conjunction with aqueous humour tap (during diagnostic work-up). A single measurement of the neutrophil blood count was used for each patient, except for two patients that had multiple measurements on the same day (patient 1, two measurements 2 minutes interval [4.16-4.23×10^9^/L] and the second patient, three measurements in a 1.5 hour time frame [3.03-3.19×10^9^/L]), which resulted in 114 samples.

The neutrophil count data for the treatment-naive North-American patient cohort (n=67, cohort 3) was determined in whole blood samples using the Sysmex XN-3000 automated haematology analyser (Sysmex Corporation, Kobe, Japan). The cumulative hazard rates were analysed using the *coxph()* function and *ggforest()* functions from the *survival* and *survminer* R packages. To determine the “best split” in neutrophil count, we iteratively estimated the maximum of the standardized log-rank statistics using *surv_cutpoint()* function of the *survminer* R package with the minimal proportion of observations per group parameter *minprop* ranging from 0.1-0.49.

### Statistical analysis

All statistical analyses were done in R version 4.0.3 (2020-10-10). The full reproducible code, raw data, and metadata is available at https://doi.org/10.34894/QR1VFZ, *DataverseNL*. A conservative cut-off for relative fluorescence units (RFUs) of 200 was used for detection of proteins (9). Aptamers with a mean RFU<200 in all disease groups were removed, leaving 938 aptamers. Two outlier samples were identified by principal component analysis (Figure 1B) and removed. Data for 78 samples was subjected to quantile normalization using the R package “preprocessCore” with the function *normalize*.*quantiles()*, and subsequently subjected to Box-Cox transformation with the *preProcess()* function and method parameter including “center”, “scale”,”BoxCox”, and “nzv”, which mean-centered and scaled the data, and removed data with near zero-variance (n=2 aptamers removed). Differential expression analysis was conducted on 936 aptamers using a likelihood ratio function adjusting for age and sex by adding these as covariates to the linear models. The *qvalue* R package was used for false discovery rate (FDR) estimation and q<0.05 was considered statistically significantly. Pathway enrichment analysis was conducted using the *ClusterProfiler* package (11) and *WikiPathways* (12).

## Data Availability

A link to the full reproducible code and data used in this study can be found in the Methods.

https://doi.org/10.34894/QR1VFZ

## Study approval

This study was conducted in compliance with the Declaration of Helsinki. Ethical approval was obtained from the Medical Ethical Research Committee of the University Medical Center Utrecht. All patients signed written informed consent before participation.

## Author contributions

JK, JHB, TR, conceived and designed studies. SH, RW and FV conducted patient recruitment, sample preparation, and JHB, NS, MY collected clinical data and conducted blood analysis. AH and KC contributed to the SOMAscan analysis. JK, IH, SH, JD, analyzed data. JK wrote the manuscript. All authors approved the manuscript.

## Acknowledgements

Astrazeneca and Uitzicht (#2014-4) supported the work presented. Currently, TR is an employee of Abbvie where he holds stock. TR had no part in the design and interpretation of the study results after he started at Abbvie. AMH is an employee of AstraZeneca and holds stock/shares in AstraZeneca PLC. K.A.C. was an employee of AstraZeneca at the time that this analysis was conducted and is currently an employee of Regeneron Pharmaceuticals.

---

**Supplemental Figure 1.**
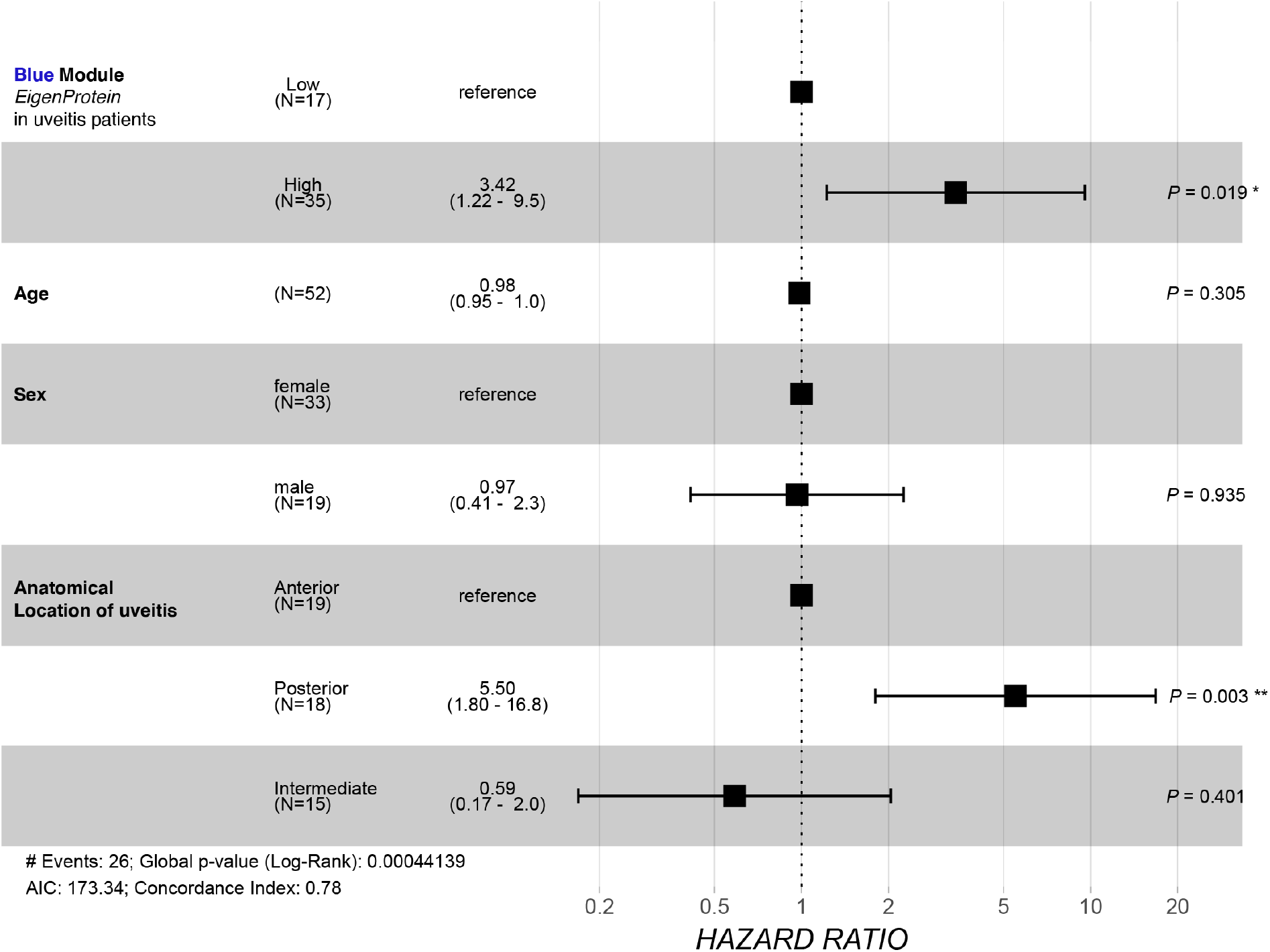
Multivariate Cox model (forest plot) of the proportional hazard for patients of cohort 1 stratified by the levels of the blue module (EigenProtein low group versus EigenProtein high group). For covariates age, sex, and anatomical location of uveitis, the hazard ratio (HR) and the 95% confidence intervals of the HR are displayed.

**Supplemental Figure 2.**
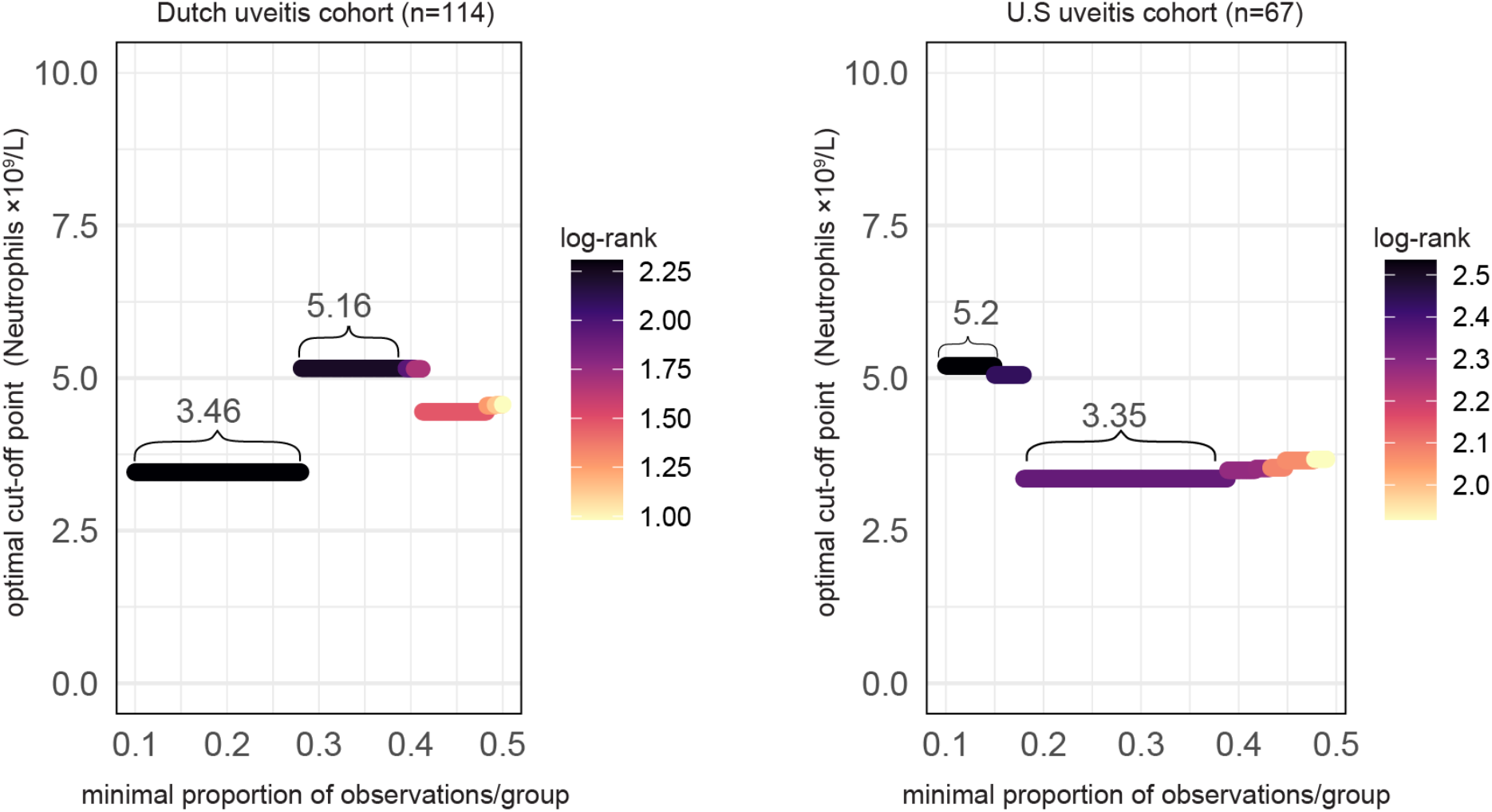
The assessment of the predictive power of neutrophil blood count split points from the Dutch and North-American cohorts for immunomodulatory therapy determined by a maximally selected rank statistic.

**Supplemental Figure 3.**
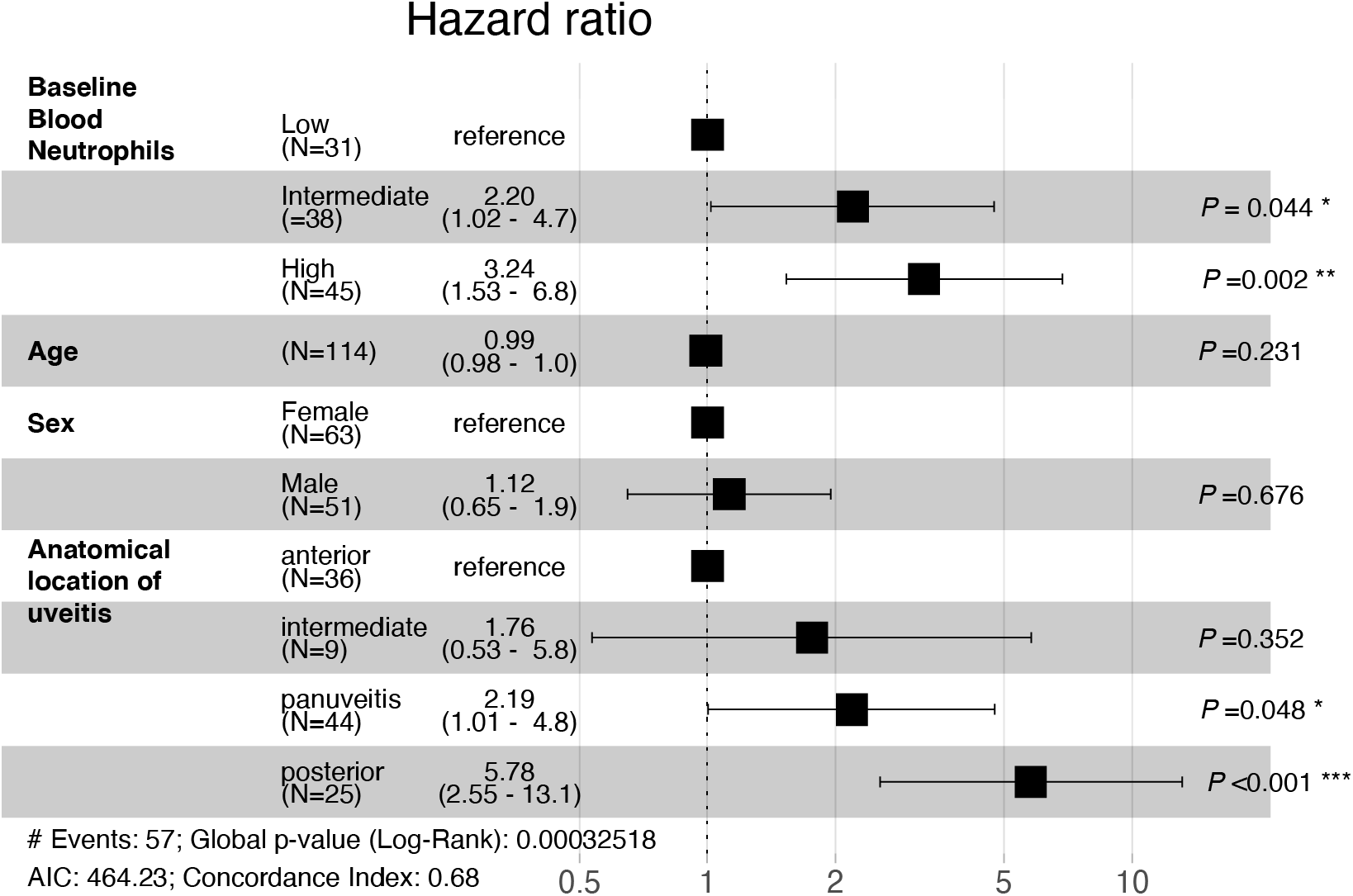
Forest plot of the multivariate Cox model for systemic immunomodulatory therapy during follow-up adjusted for age, sex, and anatomical location of uveitis for cohort 2 (n=114). Patient stratified by the baseline blood neutrophil count (proxy for the blue module); “low” (≤3.5×10^9^/L, n=31), “intermediate” (>3.5×10^9^/L and ≤5.2×10^9^/L, n=38), and “high” (>5.2×10^9^/L, n=45).

**Supplemental Figure 4.**
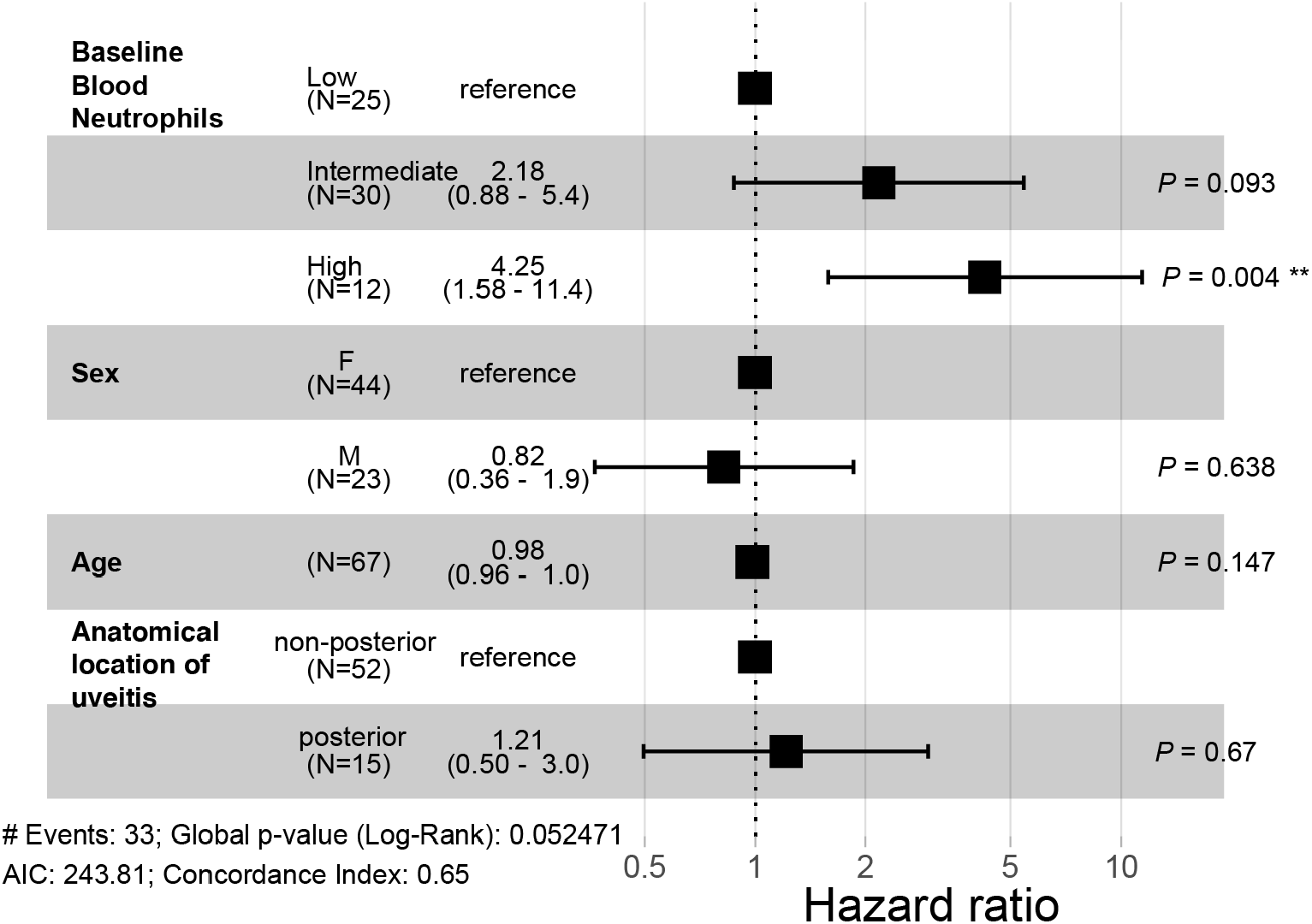
Forest plot of the multivariate Cox model for systemic immunomodulatory therapy during follow-up adjusted for age, sex, and anatomical location of uveitis for cohort 3 (n=67). Patient stratified by the baseline blood neutrophil count (proxy for the blue module); “low” (≤3.5×10^9^/L, n=25), “intermediate” (>3.5×10^9^/L and ≤5.2×10^9^/L, n=30), and “high” (>5.2×10^9^/L, n=12).

